# Mass drug administration campaigns: Comparing two approaches for schistosomiasis and soil-transmitted helminths prevention and control in selected southern Malawi districts

**DOI:** 10.1101/2022.03.21.22272679

**Authors:** Peter Makaula, Sekeleghe Amos Kayuni, Kondwani Chidzammbuyo Mamba, Grace Bongololo, Mathias Funsanani, Lazarus Tito Juziwelo, Janelisa Musaya, Peter Furu

## Abstract

Preventive chemotherapy using mass drug administration (MDA) is one of the key interventions recommended by WHO, to control neglected tropical diseases. In Malawi, health workers distribute anti-helminthic drugs annually with most support from donors. The mean community coverage of MDA from 2018 to 2020 were high at 87% for praziquantel and 82% for albendazole, however posing a sustainability challenge once donor support diminishes. This study was conducted to compare use of the community-directed intervention (CDI) approach with the use of health workers in delivery of MDA. It was carried out in three districts, where cross-sectional, mixed-methods approach to data collection during baseline and follow-up assessments was used.

Knowledge levels were high for what schistosomiasis is (65% - 88%) and what STH are (32% - 83%); and low for what causes schistosomiasis (32% - 58%), causes of STH (7% - 37%), intermediate organisms for schistosomiasis (13% - 33%) and types of schistosomiasis (2% - 26%). At follow-up, increases in praziquantel coverage were registered in control (86% to 89%) and intervention communities (83% to 89%); decreases were recorded for control (86% to 53%) and intervention schools (79% to 59%). Assessment of the costs for implementation of the study indicated that most resources were used at community (51%), health centre (29%) and district levels (19%). The intervention arm used more resources at health centre (27%) and community levels (44%) than the control arm at 2% and 4% respectively. Health workers and community members perceived the use of the CDI approach as a good initiative and more favorable over the standard practice of delivering MDA.

The use of the CDI in delivery of MDA campaigns against schistosomiasis and STH is feasible, increases coverage and is acceptable in intervention communities. This could be a way forward addressing the sustainability concern when donor support wanes.

**Trial Registration:** PACTR202102477794401

**Author summary:** World Health Organization recommends mass drug administration (MDA) as a key control measure against neglected tropical diseases. In Malawi, community-based health workers distribute drugs for schistosomiasis and soil-transmitted helminths (STH) annually, using mostly donor support which raises concern on the programme sustainability without such support. This study compared the use of the local community people as volunteers in delivery of effective MDA against schistosomiasis and STH, defined as community-directed intervention (CDI) approach, with current standard practice of using community-based health workers. The MDA coverage in both groups was noted to be high, with community-based health workers, volunteers, community leaders and people welcoming the CDI approach as good, convenient, acceptable and satisfactory initiative. Therefore, this CDI approach is a positive and sustainable move towards successful delivery of MDA against schistosomiasis and STH in endemic and limited resource settings, using local community volunteers.

## Introduction

Neglected tropical diseases (NTDs) are a diverse group of human and zoonotic diseases whose health and economic burden fall most heavily on the poorest people and communities [1]. These include diseases such as soil-transmitted helminthiases (STH), lymphatic filariasis, onchocerciasis, schistosomiasis, dengue and rabies among the 20 listed diseases [1]. Globally at least 1.7 billion people in 185 countries are affected by NTDs and require mass or individual treatment and care to minimize the burden. Of these, 1.1 billion (65%) are in low- and middle-income countries which are affected by at least five NTDs [1]. The World Health Organization (WHO) road map for prevention and control of NTDs advocates for integrated approaches in order to achieve country specific targets through cross-cutting activities that intersect multiple diseases, in order to attain the universal health coverage and sustainable development goals. Preventive chemotherapy using mass drug administration (MDA), case-management, vector management, environmental sanitation measures and health promotion are the key interventions for control of NTDs [2]. MDA, following recommendations by the WHO, is the main strategy used to control NTDs [3-5].

Malawi is endemic for several NTDs and since 1994 studies have shown schistosomiasis and STH as of major public health impact [6]. These diseases require special emphasis due to their high public health significance and varied MDA coverage. MDA is one of the community-based programmes which is widely carried out annually to prevent and control schistosomiasis and STH [7]. During annual MDA campaigns, it is mostly the Health Surveillance Assistants (HSAs) who are community-based health workers responsible for the distribution of drugs. During MDA campaigns carried out in years, 2018 to 2020, community coverage of MDA in Malawi was high at 87% (range 51.5%-95.0%) for praziquantel and 82% (range 30.6%-92.3%) for albendazole mainly due to donor support by the Schistosomiasis Control Initiative Foundation and other partners [8]. However, the existing treatment approach represents a challenge for maintaining a high, sustainable level of coverage and uptake once donors’ support comes to an end.

Evidence points to great successes in control of helminthic NTDs achieved through use of community-based interventions such as community-directed intervention (CDI) compared to use of routine health facility based or no intervention [9-11]. This is exemplified in the case of Burkina Faso, the first country in the WHO African Region to achieve nationwide coverage with anthelminthic drugs for three major NTDs namely; lymphatic filariasis, STH and schistosomiasis. Here community-based interventions have achieved high coverage without any increase in implementation costs at district and health facility levels [12]. CDI is defined as a health intervention that is undertaken by community implementers under the direction of the community itself [9]. The approach has been used successfully to distribute vitamin A and long-lasting insecticide treated nets as well as in home management of malaria [9]. The CDI strategy has also been implemented to control schistosomiasis and STH in Cameroon [13, 14], Kenya [15, 16], Malawi [17, 18], Mali [19], Nigeria [20] and Uganda [21, 22].

To the best of our knowledge, no study has tested the use of the CDI approach to deliver MDA campaigns for control of schistosomiasis and STH in Malawi. In this project, we hypothesized that the combination of publicly prioritized MDA efforts, a successful CDI programme and a well-organized community set-up would provide synergy and increased empowerment, efficiency, coverage, health impact and sustainability for MDA. This study therefore, intended to compare the use of the CDI approach with the standard practice of using community-based health workers in delivery of MDA campaigns against schistosomiasis and STH in selected districts of southern Malawi. The selection of schistosomiasis in this study was of our interest due to the fact that its drug, praziquantel requires calculation of specific dosage before administration to a person and also due the associated adverse effects which require proper observation and management.

## Methods

Reporting of this study has been verified in accordance with the Strengthening the Reporting of Observational Studies in Epidemiology (STROBE) checklist (a guidance on how to report observational research) [23].

### Study design

This study was designed as a controlled implementation research with two arms, namely; an intervention arm – which implemented MDA campaigns at community level using the study-directed CDI approach, and a control arm – the standard practice, which implemented no project-directed MDA campaigns but relied on routine campaigns organized by the National Schistosomiasis and STH Control Programme.

### Study area, target population and sample sizes

The study was carried out in the three southern Malawi districts of Chiradzulu, Mangochi and Zomba (Figure 1).

**Figure 1:**
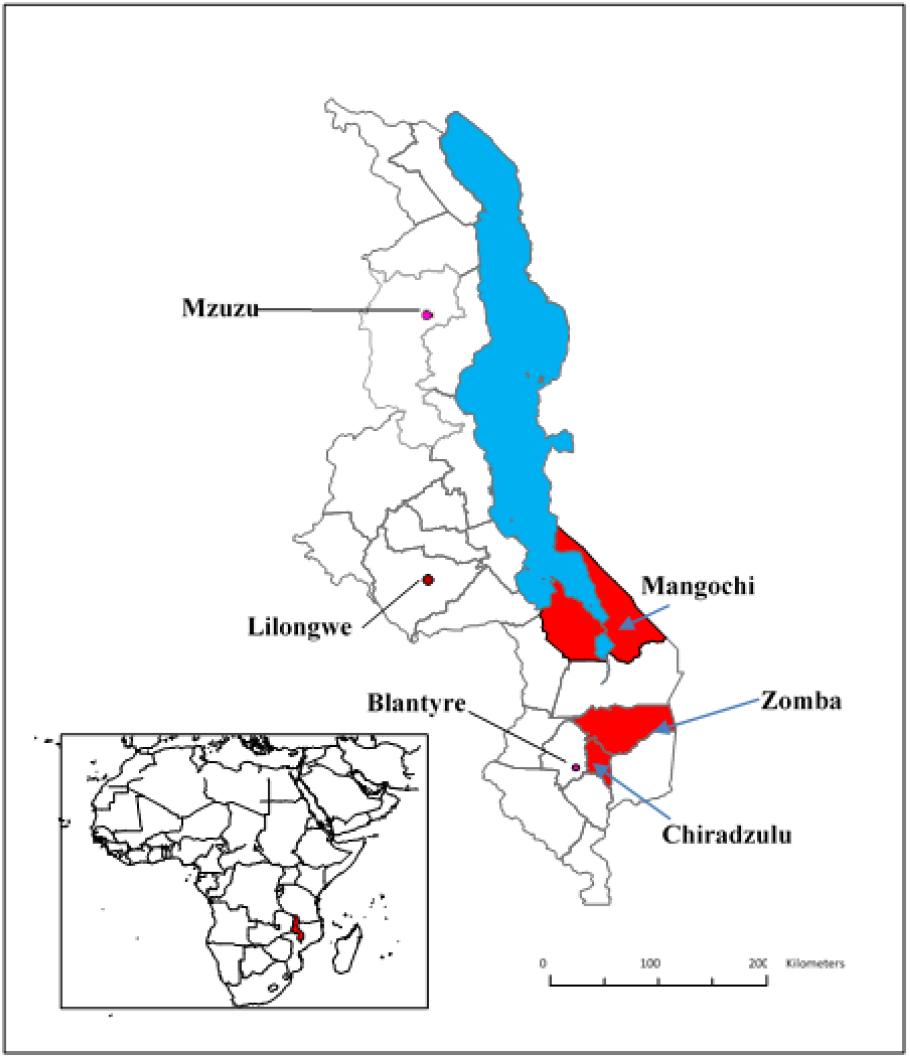
Locations of the districts of Mangochi, Zomba and Chiradzulu (in red), Lake Malawi (in blue), major cities of Mzuzu, Lilongwe and Blantyre and the location of Malawi in Africa (red in the inset) (Source: Authors’ own map [17, 18])

The study districts were selected purposively based on their level of co-endemicity with schistosomiasis and STH, and the comparative socio-economic, demographic and health indicators of the three districts [7]. In each district, four health centres and 16 villages within the catchment areas of the health centres were randomly selected to participate in the study. Each study arm was randomly allocated two health centres with eight villages in total (Table 1).

**Table 1:**
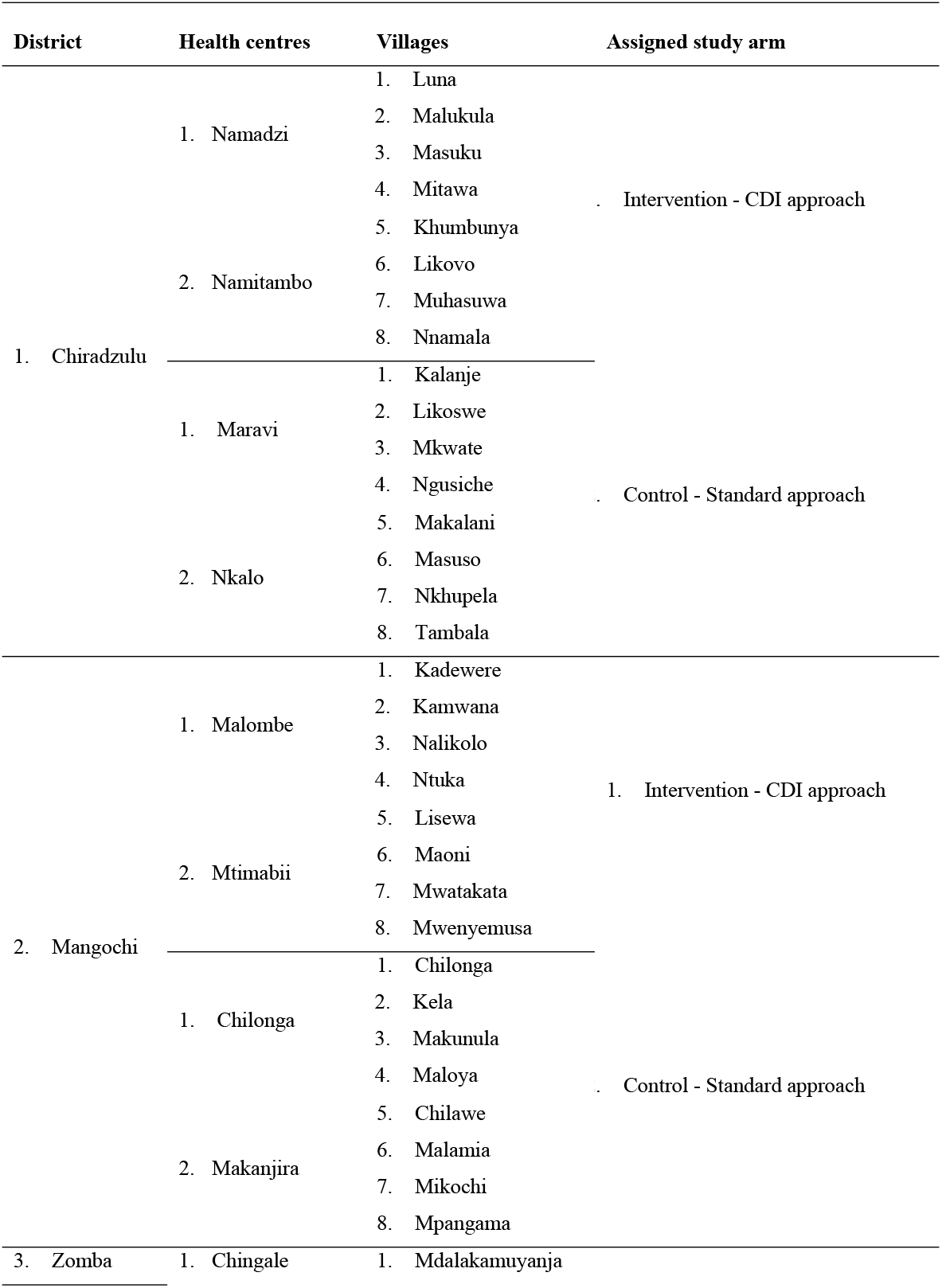
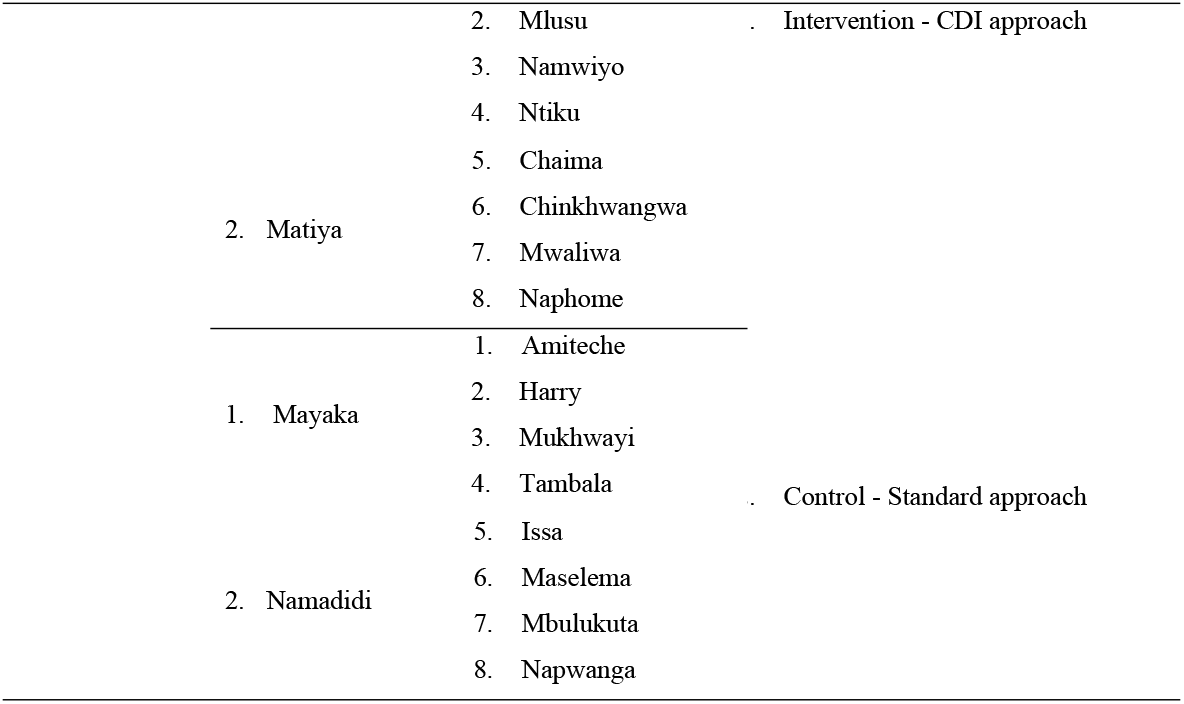
List of the involved health centres and villages according to their assigned study arms

The study population comprised the District NTD Coordinators, Pharmacy Technicians, representatives of implementation partners involved in delivery of MDA for schistosomiasis and STH, officers in-charge (clinicians or nurses), HSA, community leaders and adult community members (above age 14 years). Sampling techniques deployed at district level included purposive selection of key informants namely, the District NTD Coordinators, Pharmacy Technicians, and representatives of some of the partners involved in delivery of MDA for schistosomiasis and STH in each district for face-to-face in-depth interviews. At health centre level, clinicians or nurses in-charge or their representatives, and Senior HSA were purposively selected to participate in the study. At community level, we purposively selected responsible HSA and community leaders from the targeted villages while taking into consideration their diverse gender and roles. In addition, to obtain a varied community representation and a detailed impression of community perceptions, we randomly invited different homogenous groups of eight to ten people from selected villages to participate in a focus group discussion (FGD). Lastly, in every village a predetermined number of households were randomly selected to participate in a face-to-face questionnaire-based knowledge, attitudes and practices survey. Any household member above 14 years available during the time of the visit was invited to participate in the survey while ensuring gender balance. Data were collected in three districts comprising totals of 12 health centres and 48 villages. Table 2 summarizes the methods, purposes, levels, sample sizes and quantities of data collected in the study.

**Table 2:**
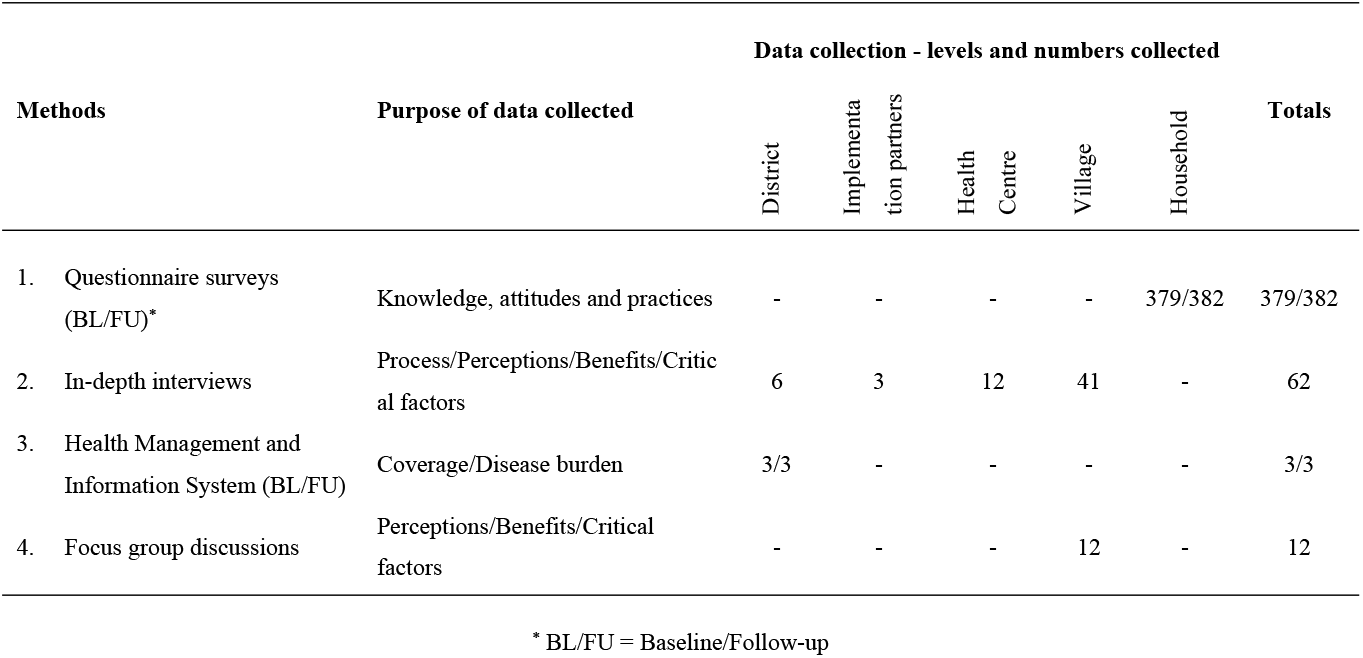
Methods, purposes, levels, sample sizes and amount of data collected in the study

### The process of using the CDI approach to deliver MDA campaigns

In the intervention arm, at community level volunteers were mobilized and sensitized on how to direct and implement MDA campaigns using the CDI approach. The CDI training guidelines and materials were adapted from the WHO [24] and the Johns Hopkins Program for International Education in Gynaecology and Obstetrics (JHPIEGO) [25]. The processes that were used to introduce CDI in the study are described as follows:

#### a) At district level

Identification of implementation partners was done, and they included Nutrition and Access to Primary Education in Chiradzulu district, Save the Children in Zomba district, and Blantyre Institute of Community Outreach in Mangochi.

The District Health Management Team selected and endorsed the District Environmental Health Officer, the District NTD Coordinator, a Community Health Nurse and a Pharmacy Technician as trainers and supervisors to implement this study. These were trained by the research team on the overall aims of the study, principles and process of the CDI approach, and on available interventions of the study. This team was responsible for training health centre-based health workers who comprised either a Medical Assistant or a nurse in charge, a Senior HSA and an HSA responsible for each of the four participating villages under the intervention arm of the study. It was the health centre-based workers who trained and supervised the community-based volunteers known as Community-Directed Distributors (CDDs).

#### b) At health centre and village levels

Trained health centre-based staff together with the HSAs responsible for the villages conducted community meetings from where CDDs or volunteers were identified (one volunteer per 200 people considering literacy and gender factors). The volunteers were trained and assigned roles as CDDs of the selected interventions with continued supervision from the health centre-based staff throughout the study implementation period.

At every stage at health centre and village levels, both the district and health centre teams participated in the training as observers to ensure quality delivery and adherence to the study protocol. In the control arm of the study, no briefing and training were offered to health staff from the corresponding health centres and villages.

#### c) Roles and responsibilities of the key players in CDI process

During the implementation of the CDI process at community level, the health services, implementation partners and the community had the following roles and responsibilities (Table 3).

**Table 3:**
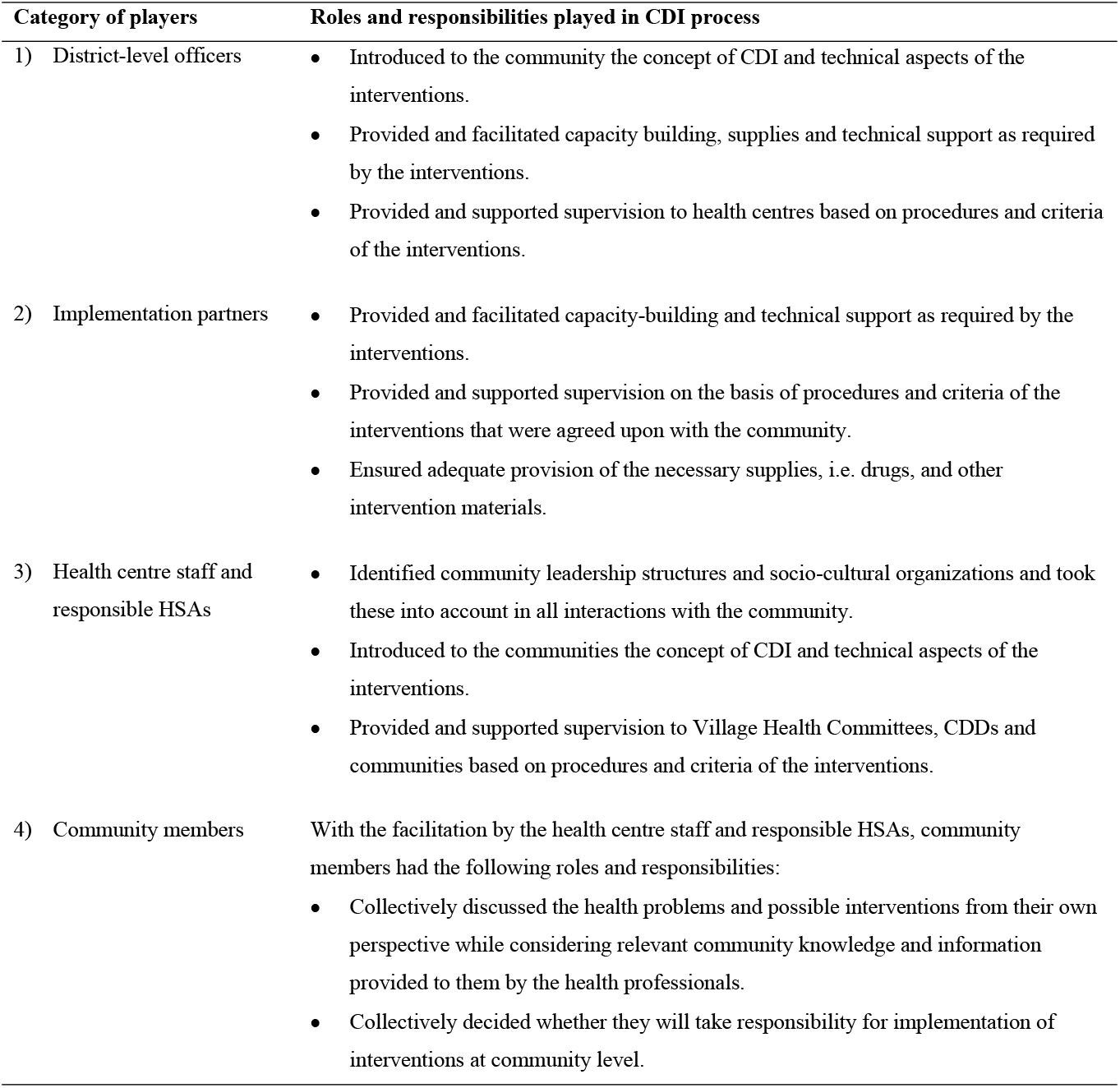
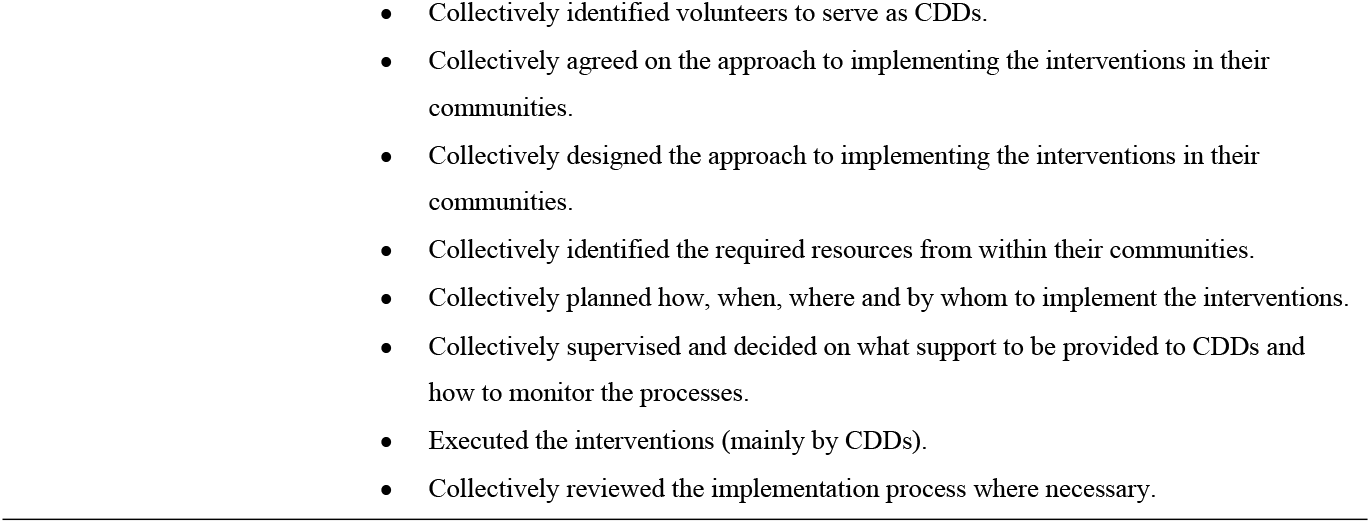
Roles and responsibilities of the key players in CDI process

| Category of players | Roles and responsibilities played in CDI process |
| --- | --- |
| 1) District-level officers | Introduced to the community the concept of CDI and technical aspects of the interventions.<br>Provided and facilitated capacity building, supplies and technical support as required by the interventions.<br>Provided and supported supervision to health centres based on procedures and criteria of the interventions. |
| 2) Implementation partners | Provided and facilitated capacity-building and technical support as required by the interventions.<br>Provided and supported supervision on the basis of procedures and criteria of the interventions that were agreed upon with the community.<br>Ensured adequate provision of the necessary supplies, i.e. drugs, and other intervention materials. |
| 3) Health centre staff and responsible HSAs | Identified community leadership structures and socio-cultural organizations and took these into account in all interactions with the community.<br>Introduced to the communities the concept of CDI and technical aspects of the interventions.<br>Provided and supported supervision to Village Health Committees, CDDs and communities based on procedures and criteria of the interventions. |
| 4) Community members | With the facilitation by the health centre staff and responsible HSAs, community members had the following roles and responsibilities:<br>Collectively discussed the health problems and possible interventions from their own perspective while considering relevant community knowledge and information provided to them by the health professionals.<br>Collectively decided whether they will take responsibility for implementation of interventions at community level. |

### Data collection

A mixed-methods approach to data collection during baseline [7] and follow-up assessments was done focusing on quantitative data for coverage and cost estimates, and qualitative data for assessing perceptions of health providers and beneficiaries, and evaluating processes regarding interventions. Cross-sectional data collection methods were used during baseline and follow-up studies where all the necessary data for the study were similarly collected from intervention and control arms during baseline and follow-up assessments. Data were collected from the involved health professionals, implementation partners, community leaders, MDA implementers and community members as beneficiaries using data collection instruments previously described [17, 18]. A questionnaire that was programmed in tablets was administered to adult household representatives at community level for determining respondents’ knowledge, attitudes and practices regarding control and prevention of schistosomiasis and STH, and delivery of MDA. Moreover, schistosomiasis and STH treatment records for years of 2019 (baseline) and 2020 (follow-up) were obtained and reviewed using checklists to establish MDA coverage data at district, health centre and village levels. Qualitative data collection instruments comprised checklists were administered to NTD Programme Coordinators and health professionals at district and health centre levels, community-based health workers and leaders at community level, to evaluate the processes used during interventions delivery and for determining perceptions of health providers and beneficiaries, benefits and critical factors. Focus group discussion guides were used to conduct group interviews with beneficiaries about their perceptions on using the interventions and benefits. All the proceedings of the key informant interviews and FGDs were recorded using digital audio recorders. Finally, document reviews were carried out in order to get an insight on the national prescription of health policy, priority health issues, costs, strategy and effectiveness of MDA delivery, availability of resources for MDA and the existing challenges and opportunities.

### Data management and analyses

Quantitative data collected through survey questionnaires was programmed in tablets using the Secure Data Kit [26]. Questionnaire and checklists data were processed and analyzed using IBM Statistical Package for Social Sciences (SPSS) software version 26. Analysis involved calculation of percentages, tabulations and frequencies to estimate MDA coverage. Furthermore, statistical significance tests using Chi Square were performed on differences in delta values (i.e. differences between baseline and follow-up) for MDA coverage for praziquantel (schistosomiasis) and albendazole (STH) between intervention and control groups. The analyses of costs and benefits data were carried out using procedures outlined by Makaula et al. (2019) [18]. Qualitative data consisted of textual and audio data, including transcripts of key informant interviews, transcripts of FGDs, field notes on observations and other intervention-specific insights, notes and reports from meetings. Transcripts were translated into English. A computer-assisted qualitative content analysis of the data using NVivo 12 for Windows (QSR International), a qualitative data analysis software programme. Data were analyzed using open coding to come up with cross-classification and retrieval of categories of texts by theme.

### Ethical considerations

The study was registered and approved by the Pan African Clinical Trial Registry (www.pactr.samrc.ac.za/) and with the number PACTR202102477794401. The study protocol including the informed consent forms were reviewed and approved by the Malawi’s National Health Sciences Research Committee (NHSRC) with the number 19/11/2443. Permissions were also sought from the respective District Councils’ health authorities and communities prior to the start of the project. Informed consent was individually obtained from every participant using local languages of Chichewa and Chiyao.

## Results

### Comparison of schistosomiasis and STH knowledge levels between intervention and control arms

As part of implementation of MDA, behavior change communication messages for schistosomiasis and STH prevention and control were also delivered in order to bring about positive changes in knowledge regarding the diseases and MDA amongst people residing in the communities. In order to assess levels of knowledge changes, the study conducted two population-based questionnaire surveys in 2020 as baseline [7] and in 2021 as follow-up for intervention and control arms.

During baseline survey, a total of 379 respondents comprising 268 (70.7%) female and 111 (29.3%) male were reached, while during follow-up a total of 382 respondents comprising 258 (67.5%) female and 124 (32.5%) male were reached. The mean age of the respondents during baseline was 40.7 years (range: 16-89) while during follow-up the mean age was 35.7 years (range: 15-83). Table 4 summarizes distribution of the demographic and socio-economic characteristics of the respondents during baseline and follow-up for the three study districts.

**Table 4:**
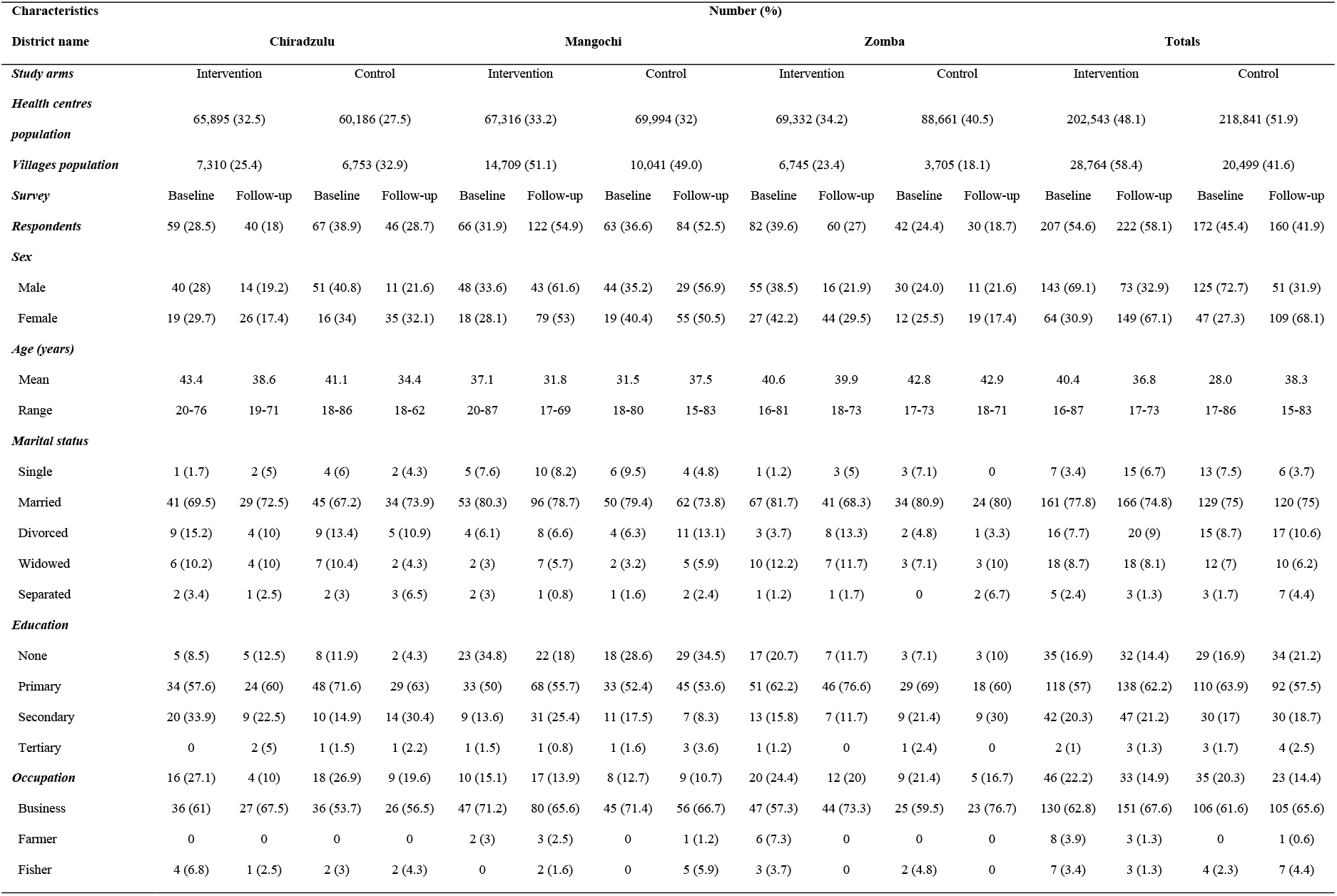
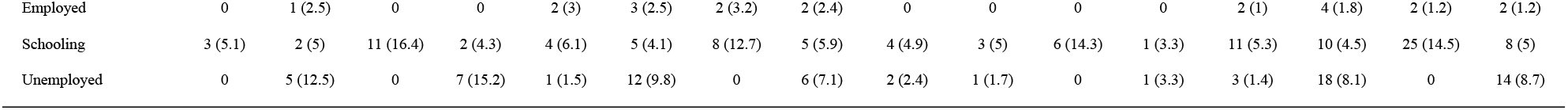
Demographic and socio-economic characteristics of survey respondents in three districts

A comparison between baseline and follow-up surveys results revealed that knowledge levels about schistosomiasis and STH varied both in intervention and control arms across the study districts. Majority of the respondents are more knowledgeable about what schistosomiasis is (range: 65% - 88%) during baseline and follow-up both in intervention and control communities. However, respondents’ knowledge on causes of schistosomiasis (range: 32% - 58%), its intermediate host (range: 13% - 33%) and its types (range: 2% - 26%) were less known or understood as few people gave the correct answers. In all the districts, the results showed that knowledge levels increased for causes of schistosomiasis (41% to 44%) and name of the intermediate organism for schistosomiasis (16% to 22%) for intervention and control arms, and types of schistosomiasis (9% to 13%) for intervention arm. Decreases in knowledge levels occurred on what schistosomiasis is (81% to 68%) for intervention and control arms; and the knowledge of types of schistosomiasis (13% to 9%) in control arm. Among the districts, Zomba (range: 7% - 88%) was highest in terms of knowledge levels, followed by Chiradzulu (range: 2% - 85%) and Mangochi (range: 7% - 71%). Figure 2 shows how the respondents answered selected knowledge-based questions about schistosomiasis during the surveys according to study arms and districts.

**Figure 2:**
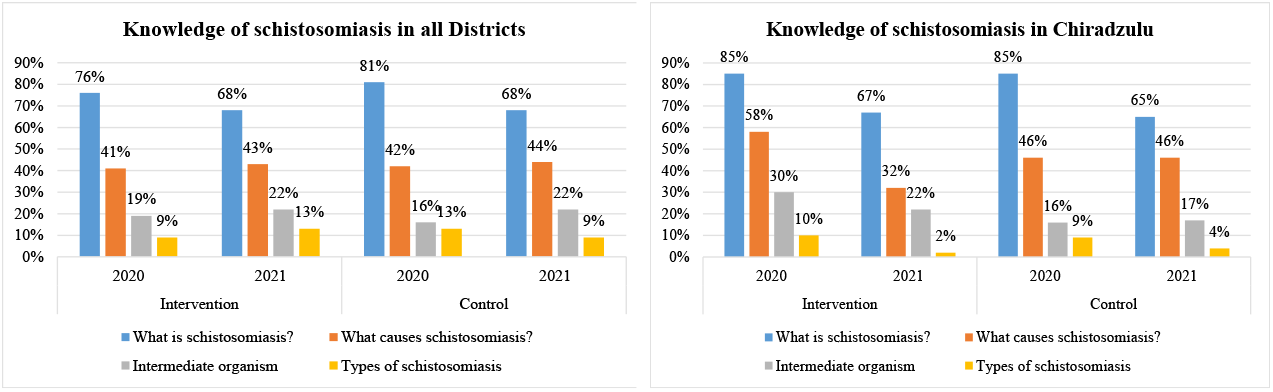
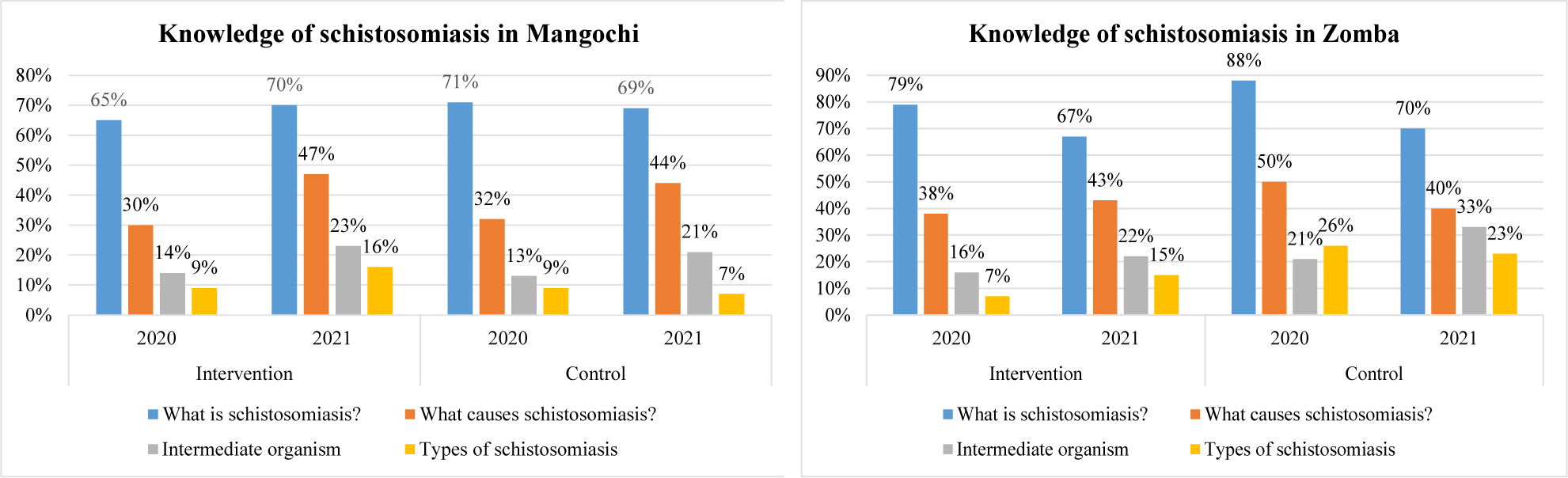
Comparison of knowledge of schistosomiasis between intervention and control arms

With regards to knowledge of STH, a comparison between baseline and follow-up results revealed that communities in intervention and control arms in the districts have varying understanding of the diseases. The surveys revealed that a majority of the respondents are knowledgeable about what STH are (range: 32% - 83%) during both baseline and follow-up, and both in intervention and control communities. However, very low knowledge levels were obtained when respondents were asked to mention what causes STH (range: 7% - 37%). In all districts, knowledge levels increased for all two indicators on what STH are (48% to 64%) and causes of STH (15% to 26%) for intervention and control arms. In the districts, Zomba (range: 21% - 83%) was highest in terms of knowledge levels, followed by Chiradzulu (range: 7% - 76%) and Mangochi (range: 9% - 50%). Chiradzulu registered decreases in knowledge of what STH are (59% to 40%) in intervention arm, causes of STH (28% to 7%) in intervention and control arms. Figure 3 shows how the respondents answered to selected knowledge-based questions about STH during the surveys according to study arms and districts.

**Figure 3:**
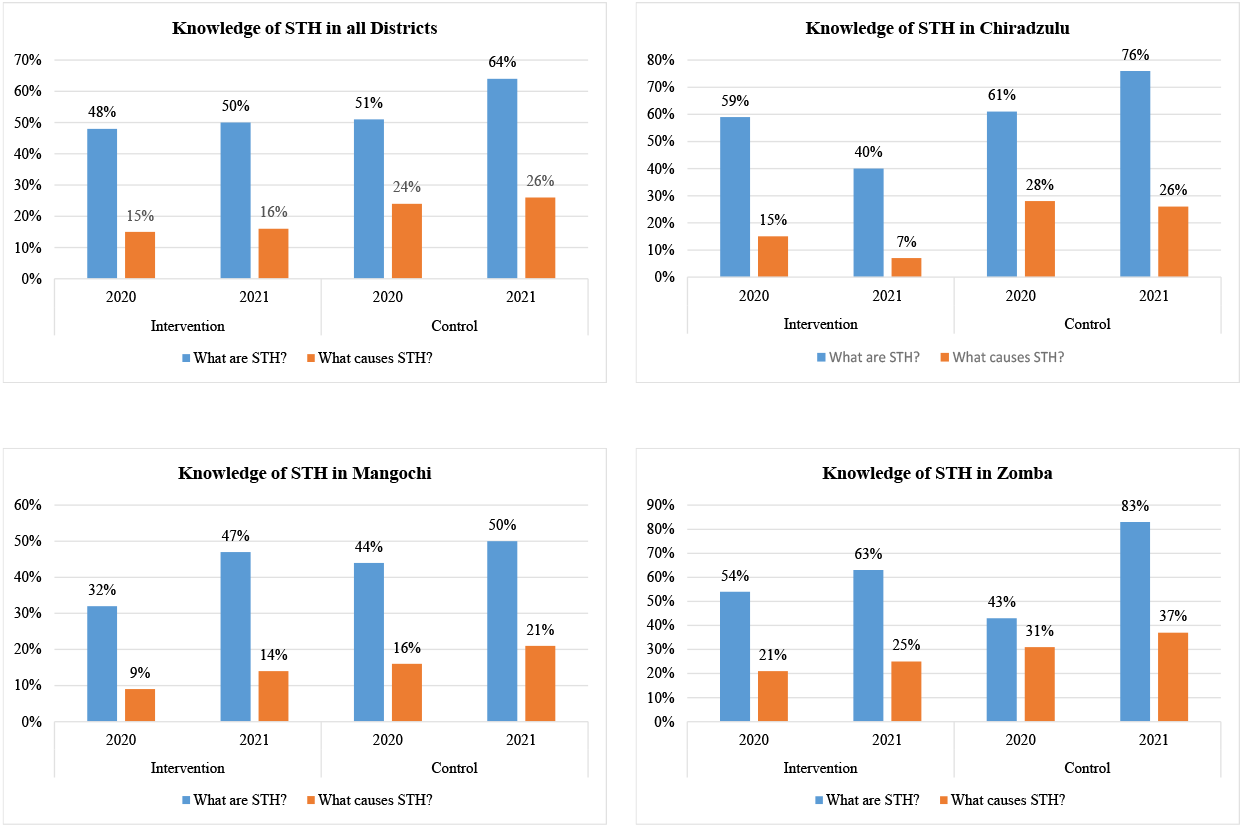
Comparison of knowledge of STH between intervention and control arms

### Comparison of MDA coverage trends for praziquantel and albendazole in study districts

The study targeted delivery of MDA campaigns using praziquantel for schistosomiasis and albendazole for STH. The MDA deliveries for these years were done by HSAs in all the districts, except for the 2020 MDA in the intervention communities which used the CDI approach by CDDs. MDA data for praziquantel and albendazole for the three years were analyzed for comparison of coverage trends.

Comparison of praziquantel coverage rates during the years revealed that all the districts registered high coverage rates for praziquantel using community-based MDA at 83% (range 73%-100%) and school-based MDA at 87% (range 75%-92%). For praziquantel community-based MDA, Chiradzulu district scored highest rates in the three years. As for praziquantel school-based MDA, Zomba district was highest in 2018 and 2019, while Mangochi district came highest in 2020. Figure 4 illustrates praziquantel coverage trends for the study districts during the three years.

**Figure 4:**
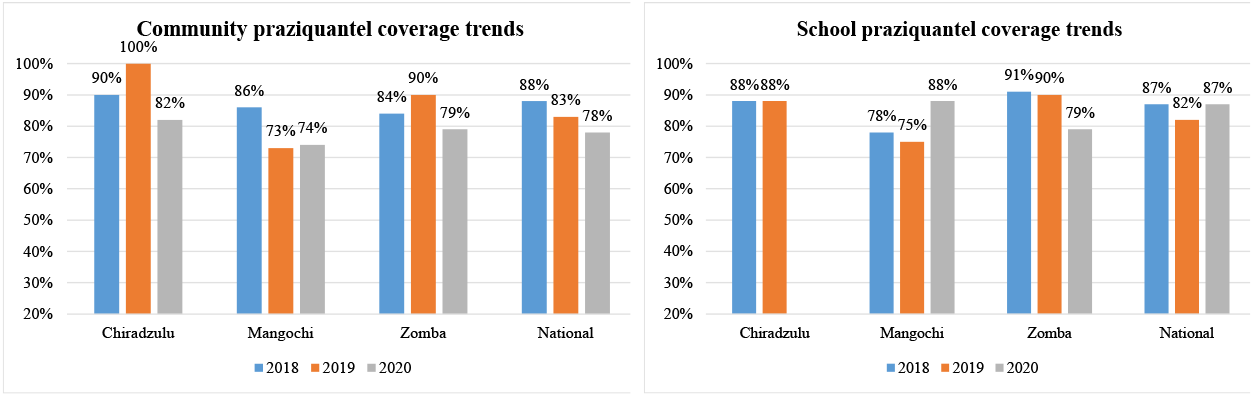
Comparison of praziquantel coverage trends in communities and schools for study districts over three years

A comparison between albendazole MDA coverage rates for 2018 and 2019 revealed that there were no differences among the districts in terms of distribution although the 2019 rates were higher than those obtained in 2018. No differences were also observed for both albendazole coverage for community and school modes of MDA deliveries. High coverage trends for community (range 71%-90%) and for schools (range 75%-92%) were observed in albendazole MDA in all the districts. In 2018 and 2019, Chiradzulu and Zomba respectively had highest albendazole coverage using the community MDA delivery approach. As for school coverage, Zomba and Chiradzulu were highest for 2018 and 2019, respectively. In 2020, there was no albendazole MDA done for communities and schools in the study districts with an exception of Mangochi which registered very low coverage rates in communities (12%) and schools (57%) due to a logistical supply chain problem. Figure 5 illustrates albendazole coverage trends for the study districts during the three years.

**Figure 5:**
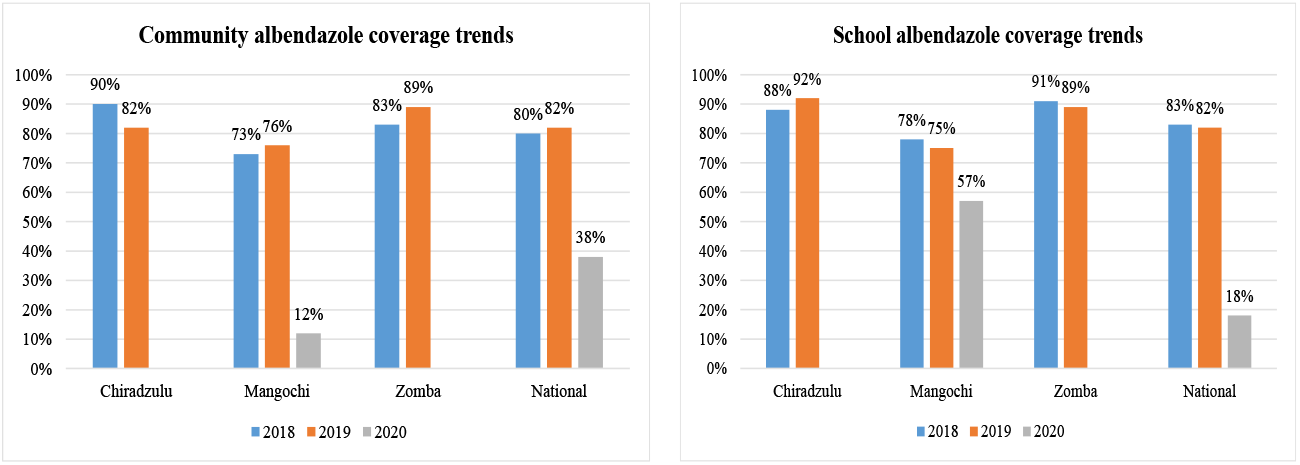
Comparison of albendazole coverage trends in communities and schools for study districts over three years

### Comparison of coverage for praziquantel in study districts by arms of study

Comparisons were made between intervention and control arms of the study for baseline and follow-up praziquantel coverage data in the three districts. Generally, in all the districts increases in coverage rates were registered in intervention (from 83% to 89%) and control (from 86% to 89%) communities, while decreases were recorded for intervention (from 79% to 59%) and control (from 86% to 53%) schools.

In Chiradzulu district, there were decreases in praziquantel MDA coverage for intervention (from 100% to 91%) and control (from 100% to 93%) communities. Despite registering high praziquantel MDA coverage rates in intervention (88%) and control (92%) schools during baseline, there was no follow-up MDA done due to schools closure brought about by the COVID-19 pandemic. There were no observed differences between coverage scores in intervention and control arms for both community and school MDA in the district.

In Mangochi district, follow-up MDA coverage rates were higher than those obtained in baseline for both community and school praziquantel MDA. Increases in coverage rates were observed in intervention (from 62% to 96%) and control (from 75% to 99%) communities; and in intervention (from 61% to 94%) and control (from 83% to 85%) schools. There were no observed differences between follow-up coverage rates in intervention and control arms and for communities and schools.

Praziquantel coverage rates in Zomba were low during follow-up than during baseline in both communities and schools. The coverage rates decreased in intervention communities (from 87% to 81%) and schools (from 87% - 83%); and control communities and schools (from 84% to 75% for both). However, there were no observed differences between coverage scores between intervention and control arms for both communities and schools meaning that there was no effect on the follow-up MDA attributable to the implementation of the study in the district. Figure 6 illustrates how the three districts generally and individually fared on praziquantel coverage for communities and schools, and according to the study arms.

**Figure 6:**
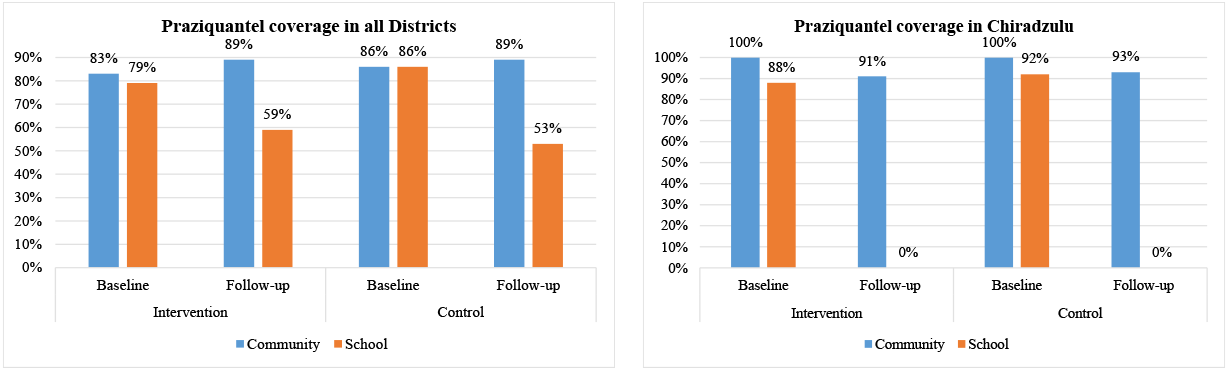
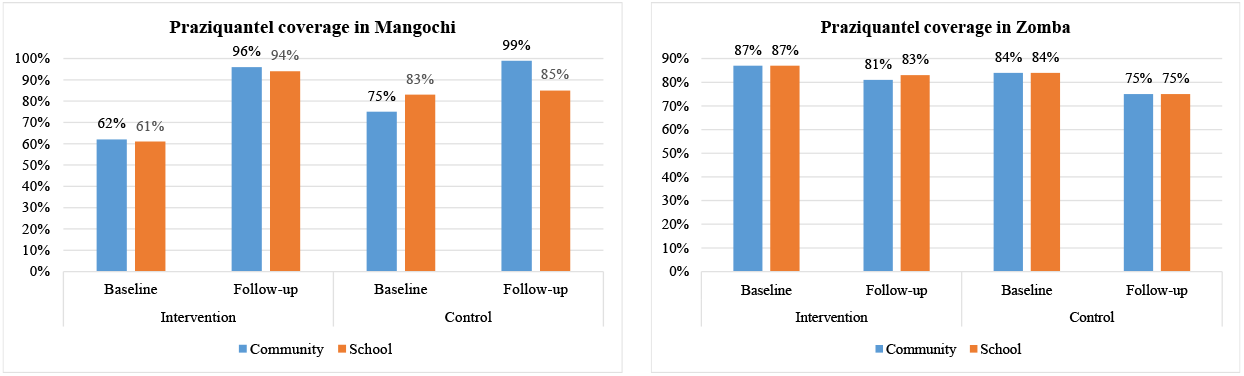
Comparison of MDA coverage for praziquantel in study districts by arms of study

There was no albendazole MDA carried out in most of the districts in 2020 due to a logistical supply chain problem with an exception of Mangochi however coverage rates were low in communities (12%) and schools (57%) to enable a comparative analysis between the study arms.

### Statistical significance of the MDA coverage difference at follow-up stage, in intervention and control groups

When differences (delta values) between baseline and follow-up coverage rates were calculated for praziquantel and albendazole in each study arm, it was observed that increases for praziquantel in communities was higher in the intervention arm (7.4%) than in the control arm (2.1%). Decreases were observed for distribution of albendazole in communities for intervention (−91.3%) and control arms (−90.4%), albendazole in schools for intervention (−58.1%) and control arms (−68.2%), and praziquantel in schools for intervention (−21.2%) and control arms (−31%). These decreases were higher in the control arm for distribution of praziquantel in schools (−31%) and albendazole in schools (−68.2%) while for distribution of albendazole in communities the decrease was higher in the intervention arm (−91.3%). Chi Square test conducted for each MDA component showed that differences in delta values in intervention and control arms were not statistically significant for all the components (Figure 7).

**Figure 7:**
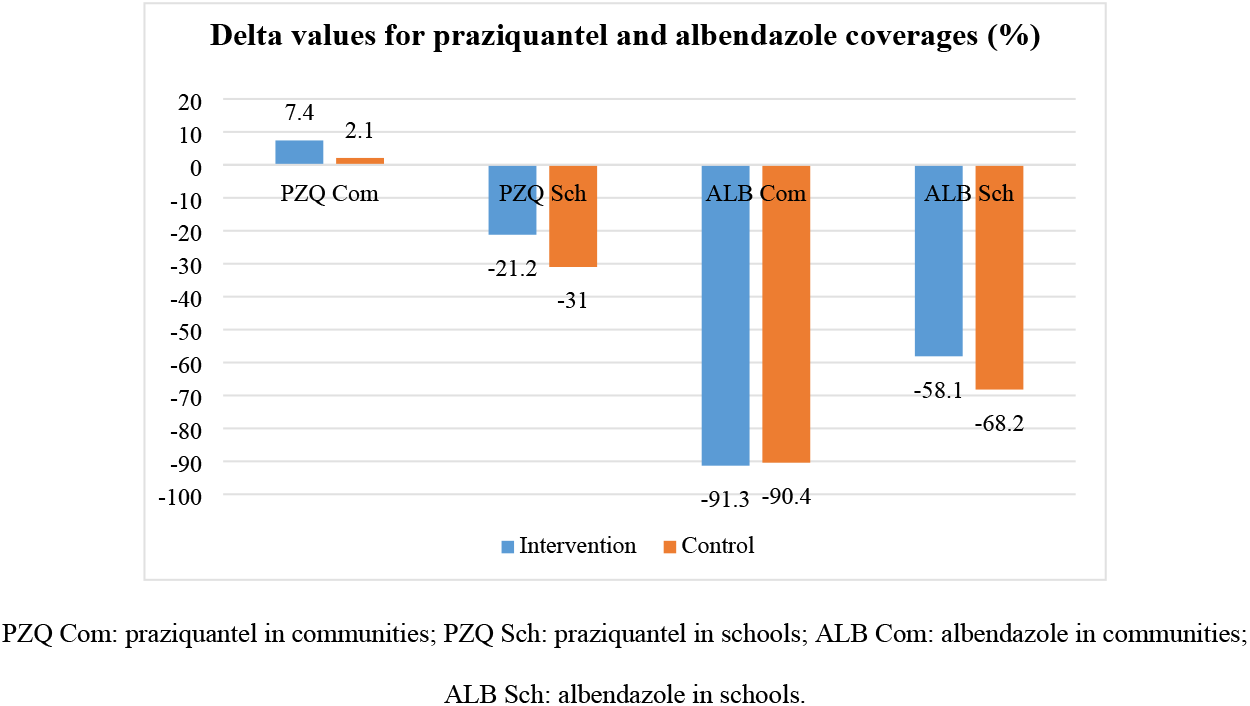
Differences in average coverage rates for praziquantel and albendazole between baseline and follow-up for intervention and control communities and schools

### Comparison of costs for using the CDI approach and the standard practice to deliver MDA campaigns

Cost data for implementation of the two approaches at three levels of study implementation: district, health centre and community were compiled and analyzed. Cost data used for comparative analysis included four main items on personnel costs of health worker and volunteer allowances during study activities, transportation, communication and other logistics such COVID-19 preventive supplies, stationery, refreshments. Out of the total amount of resources used to directly implement the selected study activities at district, health centre and community levels, most resources were used for personnel related costs (85%) followed by other logistics (9%) and transportation (5%). For both arms of the study (intervention and control), most resources were used at community level (51%) followed by health centre level (29%) and district level (19%). At both health centre and community levels, the intervention arm used more resources at 27 percent and 44 percent than the control arm at 2 percent and 4 percent respectively (Table 5).

**Table 5:** A summary of direct costs of implementing CDI and standard practice to deliver MDA in the study districts

| Cost items | | Estimated costs according to level and arm used converted to US\$* | | | | | Total US\$ for cost item (% of total) |
| --- | --- | --- | --- | --- | --- | --- | --- |
|  |  | District | Health centre |  | Community |  |  |
|  |  |  | Intervention | Control | Intervention | Control |  |
| 1. | Personnel | 1,969.73 | 4,637.13 | 435.39 | 8,314.51 | 1,421.56 | 16,778.33 (85%) |
| 2. | Transportation | 652.49 | 250.48 | - | - | - | 902.97 (5%) |
| 3. | Communication | 118.03 | 39.34 | - | - | - | 157.37 (1%) |
| 4. | Other logistics | 1,074.04 | 409.35 | - | 365.88 | - | 1,849.28 (9%) |
| Totals for level/arm (% of total) |  | 3,814.29 (19%) | 5,336.30 (27%) | 435.39 (2%) | 8,680.40 (44%) | 1,421.56 (7%) | 19,687.95 (100%) |

\*Average exchange rate: 1 United States Dollar = 762.54 Malawi Kwacha

### Perceptions of health workers and community members on the use of the CDI approach in MDA delivery

The study sought to get the perceptions of the health workers as providers and community members as beneficiaries in the intervention arm on the use CDDs in delivery of MDA. The health providers at district and facility levels perceived the use of the CDI approach to deliver MDA as a welcomed and a good initiative.

> *“I think drugs should be administered by the communities themselves, because these people reside in the same area and they know their fellow community members very well as compared to health workers.”* - A male health worker, Chiradzulu.

On the other hand, community leaders and members viewed the CDI approach convenient because people were easily accessing drugs right in their homes.

> *“It is a good development, largely because the community feels free to talk with the volunteers because they are part of them unlike the HSAs who are like strangers in the community. For example, on language they are free to communicate in their own language and can understand each other and can also trust them very easily knowing that the volunteer cannot bring something to the village to kill them.”* - KII Community leader, Mangochi.

These sentiments were shared by the CDDs who expressed satisfaction with their roles.

> *“It’s much easier for people to get medication from us because I come from the same village. People may spent more money when they go to the health centre while as within the village people will get services while doing their work at home.”* – KII female CDD, Chiradzulu.
>
> *“*[…] *there were things that we didn’t know, such as bilharzia and intestinal worm symptoms. We thought urinating blood was the only symptoms than others we have learnt.”* – KII Male CDD, Zomba.

## Discussion

The results of this study have revealed that knowledge levels in relation to schistosomiasis and STH varied disproportionately according to disease aspects asked about for both intervention and control arms across the districts. During MDA campaigns, health education messages were not comprehensively packaged and delivered in both study arms. This may have challenged the people in fully understanding these diseases, and for them to appreciate the importance of MDA and other control measures. The findings agree with other studies done in Malawi [7], Philippines [27], Nigeria [28], Egypt [29], Cameroon [30], Papua New Guinea [31] and Turkey [32] demonstrating that despite implementation of numerous activities towards the control of NTDs, there was little sensitization of the general public in order to increase awareness of the diseases. The need to engage the public in schistosomiasis and STH prevention and control activities has become imperative in the context of morbidity reduction through MDA delivery and community participation. An increase in the awareness of community members regarding schistosomiasis and STH is important for them to play a central role and to engage actively in the prevention and control of these diseases [7].

The study’s findings have also revealed that the trends for MDA coverage rates for praziquantel and albendazole from 2018 to 2020 were high in the three study districts. The results have shown that there were no differences observed between coverage trends for praziquantel and albendazole, nor for communities and schools. These high rates trends obtained in the districts during the three years were similar to the national average coverage rates indicative that as a country Malawi performance is satisfactory and probably on the right path towards the goal of reducing schistosomiasis and STH as public health problems [7, 33]. There are though some variations observed on how the individual districts and study arms performed. These variations observed between intervention and control communities in follow-up when compared to those obtained in baseline mean that other district-specific organizational factors than those attributed to implementation of the study might have influenced the results. These high MDA coverage rates are similar to what other recent studies carried out in Philippines [27], Ghana [34], Cote d’Ivoire, Kenya, Mozambique, Niger and Tanzania [35, 36], Sierra Leone [37], Togo [38] and Zanzibar [39]. A spatiotemporal modelling review [40] have also reported that schistosomiasis prevalence in sub-Saharan Africa has decreased considerably, most likely explained by the scale-up of preventive chemotherapy.

The study findings have furthermore shown that during follow-up evaluation there were no observed differences in praziquantel and albendazole MDA coverage rates between intervention and control arms for both communities and schools. It was noted that the delta values had increases only for distribution of praziquantel in communities for the intervention and control arms of the study, while various degrees of decreases occurred for praziquantel in schools in the intervention and control arms, and for all distributions of albendazole in schools and communities for both intervention and control arms. These observed decreases are contrary to what was hypothesized in the study for increased coverage rates in the intervention communities as compared to control communities. These findings demonstrate that there was no effect attributable to the implementation of the study apart from demonstrating the feasibility of using the CDI approach to deliver MDA campaigns in the study districts. We believe that since this was first time the CDI approach was used in Malawi for delivery of MDA for praziquantel and albendazole, there is a likelihood that the coverage rates may increase with subsequent implementation because CDDs will now be familiar with the process. A major contributing factor for variations in coverage rates that were obtained in this study was due to emergence of the COVID-19 pandemic that was a stress factor for the health system and might have influenced compliance and uptake of MDA in both intervention and control communities. The decreases related to distribution of albendazole were mostly due to non-availability of drug stocks at national level during the 2020 MDA campaign and the few that were treated used districts’ drug balances from the previous 2019 MDA campaign. Previous studies which were carried out elsewhere in Kenya [15, 16], Malawi [18], Mali [19], Nigeria [20] and Uganda [21, 22] reported higher treatment coverage rates in areas where CDI was used than where it was not used. We attribute the slightly different findings obtained in this study to the fact that the study was conducted under the COVID-19 pandemic condition which might also have negatively influenced the results due to misinformation associated to distribution of drugs or vaccine in communities.

A comparative analysis of the cost data for implementation of the two approaches has revealed that the intervention arm used more resources (about eight times) than the control arm. More than half of the total direct resources were used at the community level followed by health centre and district levels. For the cost items, most of the total resources used at all levels for both arms went to personnel related costs followed by other logistics and transportation costs. Generally, the implementation of the study incurred more resources than originally planned due to the emergence of the COVID-19 pandemic. The study incurred additional costs related to procurement of COVID-19 personal protective equipment for use by the researchers, participants and trainees in the various planned activities. Duplication of activities were also made in some instances to minimize the number of participants in sessions with additional transport costs and staff time. The considerable high direct costs of implementing the CDI approach may be considered a disincentive in the short term, however, if considered as an investment, it may become cheaper in the long term due to the cumulative effect of disability-adjusted life years gained through resultant improved health services or due to the development of community capacity to handle its own health challenges. This is truer when the intervention is juxtaposed with the standard practice where it is mostly dependent on donor support which cannot always be guaranteed. With the study mostly registering no observable differences in scores between the intervention and control arms, it would be reasonable to postulate that the CDI approach is equally capable of producing the same results as the standard approach. The CDI approach is implementable at a lower cost in the long term than the standard approach if the once off cost of implementing the initial investment activities like training of communities to deal with their own health issues. The costs of indirect leveraged mostly in-kind contributions related to administrative, logistical, personnel, supplies, drugs, infrastructure and technical expertise made by the participating institutions towards implementation of the study have not been included in this analysis. Indirect contributions are estimated to have covered 50% of the overall costs of the study. If these indirect contributions were to be included in the determination of the overall costs then it would not be cost effective to use the CDI approach to deliver MDA campaigns against schistosomiasis and STH.

The main challenge related to implementation of the study were disruptive delays due to the COVID-19 pandemic, which challenged the original intended timing of the study as well as the budget. This included the experienced need for appropriate timing and communication with District Health authorities, who had to focus more on urgent COVID-19 matters than on schistosomiasis and STH control. There was also a delay experienced by the National Schistosomiasis and STH Control Programme in delivery of drugs for the 2020 MDA campaign in all the districts. It is encouraging though that in this study both the health workers as providers at district and facility levels and community members as beneficiaries perceived the use of the CDI approach to deliver MDA as a welcomed and a good initiative. These sentiments and perceptions are similar to those also expressed in other studies within the Sub-Sahara African region [14, 15, 18-22]. Implementation of the CDI approach to deliver MDA campaigns for prevention and control of schistosomiasis and STH can help in addressing a service delivery gap thereby making the interventions more accessible for those in need. As a recommendation for future related research in Malawi, there is a need to explore more involvement and empowerment of community members in implementation of integrated NTDs control interventions such as health information, education and communication, as well as snail control and water, sanitation and hygiene. To enhance the coverage and sustainability of the MDA campaigns, a scaling-up of use of the CDI approach in MDA delivery may benefit from experiences and use of the ExpandNet/WHO resources through *“deliberate efforts to increase the impact of successfully tested health innovations so as to benefit more people and to foster policy and programme development on a lasting basis”* [41].

In conclusion, the study has demonstrated the feasibility of using the CDI approach for delivery of MDA for prevention and control of schistosomiasis and STH in Malawi. This study has also shown that there were no observed major differences in MDA coverage rates between intervention and control arms despite some challenges encountered during implementation. The findings have revealed existence of gaps in health education messages regarding control of schistosomiasis and STH. Although it was more costly to implement the interventions in the short term, the long term benefits of using the CDI approach in delivery of MDA outweigh the investments, and both the health providers and beneficiaries perceived the interventions as good and welcome. This could be a way forward addressing the sustainability concern for schistosomiasis and STH control when donor support wanes.

## Data Availability

The datasets generated and/or analyzed during the current study are not publicly available because participants did not grant permission for public sharing of the data in our informed consent process, approved by Malawi National Health Sciences Research Committee but are available from the corresponding author on reasonable request.

## Declarations

### Ethics and consent to participate

In this study, all methods were carried out in accordance with relevant guidelines and regulations. The study protocol, including the informed consent forms were reviewed and approved by the Malawi National Health Sciences Research Committee (NHSRC) (Approval #2443) prior to the commencement of the study. Informed consent was individually obtained from every participant at the very beginning before proceeding with their involvement after being explained to clearly about the study objectives and aims using local languages of Chichewa and Chiyao used in the study areas. For participants who were literate, a written consent was obtained while for those illiterate a written informed consent to participate was obtained through their legally authorized representatives (literate family members) instead. Similarly for the study participants who were children (under 16 years of age), consent to participate was collected from their parents or guardians on their behalf. All the research instruments that were used at community level were also translated to local languages.

### Consent for publication

Not applicable. The manuscript does not contain any individual person’s data in any form.

### Availability of data and materials

The datasets generated and/or analyzed during the current study are not publicly available because participants did not grant permission for public sharing of the data in our informed consent process, approved by Malawi National Health Sciences Research Committee but available from the corresponding author on reasonable request.

### Related manuscripts

The authors declare that they do not have a related or duplicate manuscript under consideration (or accepted) for publication elsewhere.

### Authors’ contributions

PM and PF conceived the project. PM, SAK, KCM, GB, MF, LTJ, JM and PF designed the study and the tools, conducted selection and preparation of study areas, trained and supervised data collectors. PM, SAK, KCM, GB, MF, JM, LTJ and PF analyzed and interpreted the data. All authors participated in writing and approved the final manuscript.

## Acknowledgements

The authors would like to acknowledge the Director of Preventive Health Services in Ministry of Health, Dr Storn Kabuluzi, Manager for Onchocerciasis Programme in Ministry of Health, Mr Laston Sitima, Directors of Health and Social Services and staff at the District Health Offices, health centres and implementation partners in Chiradzulu, Mangochi and Zomba district councils for their involvement in this study. The study would not have been possible without the contribution of community members who willingly participated and provided information. The authors are grateful to Mr Innocent Mvula, Mr Noel Zondola (deceased), Mr Daniel Njoka-Mwanza, Mr Austin Zgambo, Mr Moses Ngwira, Mr Mustapha Maulidi and Mr Mwai Chipeta for their support rendered during the implementation of the study.

## References

1. World Health Organization. Working to overcome the global impact of Neglected Tropical Diseases. First WHO Report on Neglected Tropical Diseases. Geneva, update 2011; https://apps.who.int/iris/handle/10665/44778.

2. World Health Organization. Ending the neglect to attain the Sustainable Development Goals: a road map for neglected tropical diseases 2021–2030. World Health Organization, Geneva, 2020; https://www.who.int/neglecteddiseases/WHONTD-roadmap-2030/en/.

3. World Health Organization. Sustaining the drive to overcome the global impact of Neglected Tropical Diseases. Second WHO report on Neglected Tropical Diseases. Geneva, 2013; http://www.who.int/iris/bitstream/10665/77950/1/9789241564540_eng.pdf.

4. World Health Organization. Preventive chemotherapy in human helminthiasis. Coordinated use of anthelminthic drugs in control interventions: a manual for health professionals and programme managers. Geneva, 2006. https://apps.who.int./iris/handle/10665/43545.

5. Montresor A, Crompton DWT, Gyorkos TW, Savioli L. Helminth control in school age children: a guide for managers of control programmes. World Health Organization. Geneva, 2016. https://www.who.int/neglected_diseases/resources/9789241548267/en/.

6. Makaula P, Sadalaki JR, Muula AS, Kayuni S, Jemu SK, Bloch P: Schistosomiasis in Malawi: a systematic review. Parasit Vectors. 2014; 7:570. doi:10.1186/s13071-014-0570-y.

7. Makaula P, Kayuni SA, Mamba KC, Bongololo G, Funsanani M, Musaya J, et al. An assessment of implementation and effectiveness of mass drug administration for prevention and control of schistosomiasis and soil-transmitted helminths in selected southern Malawi districts. BMC Health Serv Res. 2022 [in review].

8. Ministry of Health and Population, National Schisosomiasis and STH Control Programme. Reports of 2018 - 2020 MDA Campaigns. Lilongwe, Malawi.

9. Special Programme for Research and Training in Tropical Diseases. Community-directed interventions for major health problems in Africa: a multi-country study final report. World Health Organization, Geneva, Switzerland. 2008. http://www.who.int/tdr/publications/documents/cdi_report_08.pdf.

10. Amazigo UV, Leak SGA, Zoure HGM, Njepuome N, Lusamba-Dikassa P. Community-driven interventions can revolutionise control of neglected tropical diseases. Trends in Parasitology 2012. 28(6):231–238. http://dx.doi.org/j.pt.2012.03.003.

11. Burnim M, Ivy JA, King CH. Systematic review of community-based, school-based, and combined delivery modes for reaching school-aged children in mass drug administration programs for schistosomiasis. PLoS Negl Trop Dis 2017. 11(10): e0006043. https://doi.org/10.1371/journal.pntd.0006043.

12. Koukounar A, Gabrielli AF, Toura S, Bosqua OA, Zhang Y, Sellin B, et al. Schistosoma haemotobium infection and morbidity before and after large-scale administration of praziquantel in Burkina Faso. J Infect Dis 2007, 196(5):659.

13. Kamga G, Dissak-Delon FN, Nana-Djeunga HC, Biholong BD, Ghogomu SM, Souopgui J, et al. Audit of the community-directed treatment with ivermectin (CDTI) for onchocerciasis and factors associated with adherence in three regions of Cameroon. Parasit Vectors 2018, 11:356 https://doi.org/10.1186/s13071-018-2944-z.

14. Dissak-Delon FN, Kamga G, Humblet PC, Robert A, Souopgui J, Kamgno J, et al. Do Communities Really “Direct” in Community-Directed Interventions? A Qualitative Assessment of Beneficiaries’ Perceptions at 20 years of Community Directed Treatment with Ivermectin in Cameroon. Trop Med Infect Dis 2019, 4:105.

15. Mwinzi PNM, Montgomery SP, Owaga OC, Mwanje M, Muok EM, Ayisi JG, et al. Integrated community-directed intervention for schistosomiasis and soil transmitted helminths in Western Kenya – a pilot study. Parasit Vectors 2012, 5:182.

16. Odhiambo GO, Musuva RM, Odiere MR, Mwinzi PN. Experiences and perspectives of community health workers from implementing treatment for schistosomiasis using the community directed intervention strategy in an informal settlement in Kisumu City, western Kenya. BMC Public Health 2016, 16:986 DOI 10.1186/s12889-016-3662-0

17. Makaula P, Bloch P, Banda H, Mbera GB, Mangani C, de Sousa A, et al. Primary Health Care in rural Malawi – a qualitative assessment exploring the relevance of the community-directed interventions approach. BMC Health Serv Res 2012, 12:328.

18. Makaula P, Funsanani M, Mamba KC, Musaya J, Bloch P. Strengthening Primary Health Care at district-level in Malawi - determining the coverage, costs and benefits of Community-Directed Interventions. BMC Health Serv Res 2019, 19:509 https://doi.org/10.1186/s12913-019-4341-5.

19. Dabo A, Bary B, Kouriba B, Sankaré O, Doumbo O. Factors associated with coverage of Praziquantel for schistosomiasis control in the community direct intervention (CDI) approach in Mali (West Africa). Infect Dis Poverty 2013, 2:11.

20. Ajayi IO, Jegede AS, Falade CO, Sommerfeld J. Assessing resources for implementing a community directed intervention (CDI) strategy in delivering multiple health interventions in urban poor communities in Southwestern Nigeria: a qualitative study. Infect Dis Poverty 2013, 2:25.

21. Chami GF, Kontoleon AA, Bulte E, Fenwick A, Kabatereine NB, Tukahebwa EM, et al. Profiling Non-recipients of Mass Drug Administration for Schistosomiasis and Hookworm Infections: A Comprehensive Analysis of Praziquantel and Albendazole Coverage in Community-Directed Treatment in Uganda. Clinical Infectious Diseases 2016, 62(2):200–207.

22. Chami GF, Kontoleon AA, Bulte E, Fenwick A, Kabatereine NB, Edridah M, et al. Community-directed mass drug administration is undermined by status seeking in friendship networks and inadequate trust in health advice networks. Social Science & Medicine 2017, 183:37–47. http://dx.doi.org/10.1016/j.socscimed.2017.04.009.

23. Elm Ev, Altman DG, Egger M, Pocock SJ, Gøtzsche PC, Vandenbroucke JP. Strengthening the reporting of observational studies in epidemiology (STROBE) statement: guidelines for reporting observational studies. BMJ. 2007, 335(7624):806–8. https://doi.org/10.1136/bmj.39335.541782.AD PMID:17947786.

24. World Health Organization, African Programme for Onchocerciasis Control (WHO/APOC): Curriculum and Training Module on the Community-Directed Intervention (CDI) Strategy for Faculties of Medicine and Health Sciences – Trainers Handbook. 2013. WHO/APOC/MG/12.2.

25. Johns Hopkins University – JHPIEGO Programme. A Training Programme in Community-Directed Intervention to Improve Access to Essential Health Services – Training Guide. 2013; http://www.jhpiego.org/en/content/training-program-community-directed-intervention-cdi-improve-access-essential-health-service.

26. Pavluck A, Chu B, Flueckiger RM, Ottesen E. Electronic Data Capture Tools for Global Health Programs: Evolution of LINKS, an Android-, Web-Based System. PLoS Negl Trop Dis. 2014; 8(4): e2654. doi:10.1371/journal.pntd.0002654.

27. Inobaya MT, Chau TN, Ng S, MacDougall C, Olveda RM, Tallo VL et al. Mass drug administration and the sustainable control of schistosomiasis: an evaluation of treatment compliance in the rural Philippines. Parasit Vectors. 2018; 11:441 https://doi.org/10.1186/s13071-018-3022-2.

28. Olamiju OJ, Olamiju FO, Adeniran AA, Mba IC, Ukwunna CC, Okoronkwo C et al. Public Awareness and Knowledge of Neglected Tropical Diseases (NTDs) Control Activities in Abuja, Nigeria. PLoS Negl Trop Dis. 2014; 8(9): e3209. doi:10.1371/journal.pntd.0003209.

29. Elfar E, Asem N, Yousof H. The awareness of neglected tropical diseases in a sample of medical and nursing students in Cairo University, Egypt: A cross-sectional study. PLoS Negl Trop Dis. 2020; 14(11): e0008826. https://doi.org/10.1371/journal.pntd.0008826.

30. Kamga HLF, Assob NJC, Nsagha DS, Njunda AL, Njimoh DL. A community survey on the knowledge of neglected tropical diseases in Cameroon. Int J Med Biomed Res. 2012; 1(2):131–140. http://dx.doi.org/10.14194/ijmbr.128.

31. Karoke W. Neglected tropical diseases: less known subjects amongst health care professionals. J Community Med Public Health. 2018; 34 (6): 254–255.

32. Ulukanligil M. Community perception of school-based deworming program in Sanliurfa, Turkey. Am J Trop Med Hyg. 2006; 75:1063–1068.

33. Witek-McManus S, Simwanza J, Chisambi AB, Kepha S, Kamwendo Z, Mbwinja A, et al. Epidemiology of soil-transmitted helminths following sustained implementation of routine preventive chemotherapy: Demographics and baseline results of a cluster randomised trial in southern Malawi. PLoS Negl Trop Dis. 2021, 15(5): e0009292. https://doi.org/10.1371/journal.pntd.0009292.

34. Abass Q, Bedzo JY, Manortey S. Impact of Mass Drug Administration on Prevalence of Schistosomiasis in Eight Riverine Communities in the Asuogyaman District of the Eastern Region, Ghana. International Journal of Tropical Disease & Health. 2020, 41(15): 18–32. DOI: 10.9734/IJTDH/2020/v41i1530355.

35. Binder S, Campbell Jr. CH, Castleman JD, Kittur N, Kinung’hi SM, Olsen A, et al. Lessons Learned in Conducting Mass Drug Administration for Schistosomiasis Control and Measuring Coverage in an Operational Research Setting. Am J Trop Med Hyg. 2020, 103(Suppl 1): 105–113 doi:10.4269/ajtmh.19-0789.

36. King CH, Kittur N, Binder S, Campbell Jr. CH, N’Goran EK, Meite A, et al. Impact of Different Mass Drug Administration Strategies for Gaining and Sustaining Control of Schistosoma mansoni and Schistosoma haematobium Infection in Africa. Am J Trop Med Hyg. 2020, 103(Suppl 1): 14–23 doi:10.4269/ajtmh.19-0829.

37. Bah YM, Paye J, Bah MS, Conteh A, Saffa S, Tia A, et al. Schistosomiasis in School Age Children in Sierra Leone After 6 Years of Mass Drug Administration With Praziquantel. Front. Public Health 2019, 7:1. doi: 10.3389/fpubh.2019.00001.

38. Bronzan RN, Dorkenoo AM, Agbo YM, Halatoko W, Layibo Y, Adjeloh P, et al. Impact of community-based integrated mass drug administration on schistosomiasis and soil-transmitted helminth prevalence in Togo. PLoS Negl Trop Dis. 2018, 12(8): e0006551. https://doi.org10.1371/journal.pntd.0006551/.

39. Knopp S, Person B, Ame SM, Ali SM, Muhsin J, Juma S, et al. Praziquantel coverage in schools and communities targeted for the elimination of urogenital schistosomiasis in Zanzibar: a cross-sectional survey. Parasit Vectors. 2016; 9:5 DOI 10.1186/s13071-015-1244-0.

40. Kokaliaris C, Garba A, Matuska M, Bronzan RN, Colley DG, Dorkenoo AM, et al. Effect of preventive chemotherapy with praziquantel on schistosomiasis among school-aged children in sub-Saharan Africa: a spatiotemporal modelling study. Lancet Infect Dis 2022; 22: 136–49. https://doi.org/10.1016/S1473-3099(21)00090-6.

41. World Health Organization. Nine steps for developing a scaling-up strategy. World Health Organization. Geneva, 2010; ExpandNet. ISBN 978 92 4 150031 9 https://www.who.int./immunization/hpv/deliver/nine_steps_for_developing_a_scalingup_strategy_who_2010.pdf.

